# Factors influencing plasma galectin-3 concentrations in catheter-bearing hospitalized patients

**DOI:** 10.1101/2021.06.04.21258341

**Authors:** Simona Iftimie, Anna Hernández-Aguilera, Ana F. López-Azcona, Helena Castañé, Elisabet Rodríguez-Tomàs, Gerard Baiges-Gaya, Antoni Castro, Jordi Camps, Jorge Joven

## Abstract

**Introduction:** Catheters are an integral part of modern medicine although their use is not without complications. Catheter-related infection triggers a strong inflammatory reaction and has been associated with high morbidity, mortality, and healthcare costs. The clinical diagnosis of catheter-related infection is made difficult by non-specific symptoms. Investigating the alterations in biochemical parameters related to infectious and inflammatory processes in these patients constitute an active line of research. The aim of this study was to investigate factors influencing the plasma concentration of galectin-3 in catheter-bearing patients and to explore its potential usefulness as an index for catheter-related infections.

**Methods:** Circulating concentrations of galectin-3, chemokine (C-C) motif ligand 2, procalcitonin and C-reactive protein were measured in 110 patients with a central venous catheter, 165 patients with a urinary catheter, and 72 control subjects.

**Results:** Catheter-bearing patients had significantly higher concentrations of galectin-3 and the other markers than the control group. We identified chronic kidney disease as an independent determinant of plasma galectin-3 concentrations in patients with a central catheter, and serum creatinine concentration, cardiovascular disease and the number of days the catheter was indwelling as determinants in urinary catheter patients. We also found that measuring galectin-3 in urinary catheter patients with a catheter-related infection was more accurate for diagnosis than the other parameters. At galectin-3 = 15 ng/mL, sensitivity was 90%.

**Conclusion:** We conclude that measurement of galectin-3 concentration may be useful for assessing the inflammatory status of catheter-bearing patients and may contribute to the diagnosis of catheter-related infection in those with a urinary catheter.

## 1. Introduction

The use of catheters is an integral part of modern healthcare, as they provide access for sample extraction for analysis, administration of medication or parenteral nutrition, and access for hemodialysis and hemodynamic monitoring. The two most widely employed types of catheters in medicine are probably the central venous catheters (CVC) and the urinary catheters (UC). However, the use of these devices is not without complications, among which catheter-related infection (CRI) stands out, since it triggers a strong inflammatory reaction and is associated with increased morbidity, mortality, and healthcare costs [1,2]. Unfortunately, the clinical diagnosis of CRI is made difficult by symptoms such as fever, chills, and hypotension, which are not specific [3]. Therefore, alterations in biochemical parameters related to infectious and inflammatory processes must be investigated with the aim of identifying biochemical markers that are capable of diagnosing infections in catheter-bearing patients., Several studies have proposed C-reactive protein (CRP), procalcitonin, or the chemokine (C-C motif) ligand 2 (CCL2) as markers of infection. However, their usefulness varies according to the clinical setting, and that is an unresolved issue [4,5].

Galectin-3 belongs to a family of β-galactoside binding proteins with a structure that allows them to participate in functions involving cellular and molecular recognition and adhesion [6]. Each galectin contains a C-terminal domain of 130 amino acids, termed a carbohydrate recognition domain, which is responsible for binding to galactose-containing sugar moieties. Galectin-3 also possesses a N-terminal domain with a unique short end continuing into a Pro-Gly-Ala-Tyr-rich repeat motif [7]. This protein has a molecular mass of 31 kDa, is strongly expressed by macrophages, and is a modulator of multiple biological functions, such as proliferation, macrophage chemotaxis, phagocytosis, neutrophil extravasation, neutrophil migration, apoptosis, vacuole lysis after infection, fibrogenesis, and angiogenesis [8-10]. The measurement of plasma galectin-3 concentration has been suggested as an index of inflammation and fibrosis in various diseases [11-13]. Several studies report that galectin-3 is a component of the innate immune system, participating in the organism’s defence mechanisms against infection, promoting macrophage and neutrophil survival and phagocytosis [14-16]. It is also able to bind to lipopolysaccharide from various types of bacteria, protecting hosts from endotoxin shock [17]. Galectin-3 is even a bactericide against *Helicobacter pylori* and a bacteriostatic against *Streptococcus pneumoniae* [9,18].

Therefore, the aim of this study was to investigate the factors influencing the plasma concentration of galectin-3 and to explore the potential usefulness of this lectin as a biomarker for CRI in hospitalized patients bearing a CVC or a UC.

## 2. Materials and methods

### 2.1. Ethics approval

The study was conducted in accordance with the Declaration of Helsinki, and was approved by the Ethics Committee (Institutional Review Board) of the *Hospital Universitari de Sant Joan* (12proj2. 10/12/23). All the subjects provided written informed consent to participate in the study on the understanding that anonymity of data was guaranteed.

### 2.2. Participants

This was post-hoc retrospective cohort study including new objectives derived from a previous prospective, longitudinal study searching for biomarkers of catheter-related infection [19,20]. We selected 110 patients with a CVC and 165 patients with a UC, hospitalized in the Department of Internal Medicine or in the Intermediate Care Unit of our Institution. The control group consisted of 72 healthy volunteers, who participated in an epidemiological study conducted in our geographical area; the details of that study have been reported previously [21]. Those subjects had no clinical or biochemical evidence of renal insufficiency, liver disease, neoplasia or neurological disorders. The patients and controls were selected so that the different groups had the most similar possible age and gender distributions. Serum and plasma samples from all participants were stored in our Biobank at -80ºC until the time of the study. We recorded demographic data, comorbidities, other acute or chronic infections, and bacteriological and therapeutic data. We also calculated the McCabe score as an index of clinical prognosis [22] and the Charlson index as a way of categorizing the patients’ comorbidities [23]. Of the participants, 116 (42%) were hospitalized for surgery, 72 (26%) for an infectious disease, and the remaining 87 (32%) had various other clinical conditions. In patients with a CVC, the location of the catheter tip was brachial in 47 patients (43%), subclavian in 51 (46%), jugular in 9 (8%), and femoral in 3 (3%).

Patients with an acute concomitant infection (ACI) were those suffering from an infection (abdominal abscess, pneumonia, etc.) that was not related to an infected catheter. Sixty-one (22%) patients had an ACI without a CRI, 29 (11%) patients had a CRI without an ACI, and 24 (9%) patients had both infections simultaneously.

### 2.3. Biochemical analyses

The ethylene diamine tetraacetate (EDTA)-plasma concentrations of galectin-3 and CCL2, and the serum concentrations of procalcitonin were determined by enzyme-linked immunosorbent assay (R&D Systems^®^, Minneapolis, USA, Biovendor, Brno, Czech Republic, and Peprotech, London, UK, respectively). Serum CRP concentrations were measured by a high-sensitive solid phase chemiluminescent immunoassay (Immulite^®^ 2000 High Sensitivity CRP, Siemens, Munich, Germany) in an Immulite^®^ 2000 Xpi automated analyzer (Siemens). Serum creatinine concentrations were analyzed by the Jaffe method in a Cobas^®^ 8000 modular platform (Roche Diagnostics, Basel, Switzerland).

### 2.4. Statistical analyses

All calculations were made using the SPSS 24.0 statistical package (SPSS Inc., Chicago, IL, USA). Since most of the studied variables had non-Gaussian distributions, differences between any two groups were assessed by the Mann-Whitney *U* test. Qualitative data were analyzed using the χ^2^ test, and correlations by the Spearman’s ρ test. The combined effect of clinical and demographic characteristics on plasma galectin-3 concentrations was assessed by multiple regression analysis. The diagnostic accuracy of the measured biochemical variables was assessed by receiver operating characteristics (ROC) curves [24]. Results are shown as medians and interquartile ranges (IQR).

## 3. Results

Patients with a UC were significantly older and the percentage of males was higher than patients with a CVC. They more often had arterial hypertension, cardiovascular disease, chronic kidney disease, and/or treatment with antibiotics, and significantly less often had neoplasia and/or treatments with immunosuppresive drugs. The McCabe score indicated a better prognosis in UC patients, and the Charlson index was similar between groups. They did not show any significant difference in the incidence of CRI or ACI (Table 1).

**Table 1.** Demographic and clinical characteristics of the patients and the control group.

| Feature |  | Control group<br>(n = 72) | CVC<br>(n = 110) | UC<br>(n = 165) |
| --- | --- | --- | --- | --- |
| Age, years |  | 63 (61-69) | 63 (50-74) | 75 (63-81)<sup>c,f</sup> |
| Gender (male), n (%) |  | 47 (65.3) | 54 (49.1)<sup>a</sup> | 118 (71.5)<sup>f</sup> |
| Smoking, n (%) |  | 26 (36.1) | 25 (22.7)<sup>b</sup> | 24 (14.5)<sup>c</sup> |
| Alcohol intake, n (%) |  | 36 (50.0) | 15 (13.6)<sup>c</sup> | 35 (21.2)<sup>c</sup> |
| Catheter-related infection |  | NA | 23 (20.9) | 30 (18.2) |
| Acute concomitant infection |  | NA | 36 (32.7) | 49 (29.7) |
| Hypertension, n (%) |  | 5 (6.9) | 57 (51.8)<sup>c</sup> | 115 (69.7)<sup>c,e</sup> |
| Diabetes mellitus, n (%) |  | 0 | 32 (29.1) | 60 (36.4) |
| Dyslipidemia, n (%) |  | 0 | 35 (31.8) | 66 (40.0) |
| Cardiovascular disease, n (%) |  | 0 | 11 (10.0) | 37 (22.4)<sup>e</sup> |
| Chronic kidney disease, n (%) |  | 0 | 10 (9.1) | 35 (21.2)<sup>e</sup> |
| Neurological vascular disease, n (%) |  | 0 | 12 (10.9) | 12 (7.3) |
| COPD, n (%) |  | 0 | 11 (10.0) | 31 (18.8)<sup>a</sup> |
| History of neoplasia, n (%) |  | 0 | 71 (64.5) | 75 (45.5)<sup>e</sup> |
| Immunosuppressive treatment, n (%) |  | 0 | 45 (40.9) | 6 (3.6)<sup>f</sup> |
| Antibiotics 24h before the study, n (%) |  | 0 | 51 (46.4) | 141 (85.5)<sup>f</sup> |
| Charlson index | No comorbidity, n (%) | NA | 30 (27.3) | 67 (40.6) |
|  | Low comorbidity, n (%) | NA | 47 (42.7) | 54 (32.7) |
|  | High comorbidity, n (%) | NA | 33 (30.0) | 44 (26.7) |
| McCabe index | RFD, n (%) | NA | 9 (8.2) | 18 (10.9) |
|  | UFD, n (%) | NA | 28 (25.5) | 13 (7.9)<sup>f</sup> |
|  | NFD, n (%) | NA | 73 (66.4) | 134 (81.2)<sup>f</sup> |
| Days with catheter |  | NA | 22 (10-130) | 4 (2-7)<sup>f</sup> |

COPD: Chronic obstructive pulmonary disease; CVC: Central venous catheter; NA: Not applicable; NFD: Nonfatal disease; RFD: Rapidly fatal disease; UC: Urinary catheter; UFD: Ultimately fatal disease. Data are shown as medians and interquartile ranges or number of cases and percentages. <sup>a</sup>: $P < 0.05$ , <sup>b</sup>: $P < 0.01$ , <sup>c</sup>: $P < 0.001$ , with respect to the control group; <sup>d</sup>: $P < 0.05$ , <sup>e</sup>: $P < 0.01$ , <sup>f</sup>: $P < 0.001$ , with respect to the CVC patients. Statistical analyses were performed by the Mann-Whitney $U$ test (quantitative) or the $X^2$ test (qualitative).

Patients with a CVC or a UC had significantly higher concentrations of galectin-3, CRP, and CCl2 than the control group. They also showed a trend for higher procalcitonin concentrations, but differences did not reach statistical significance (Fig. 1). The bivariate analysis of the associations of the selected biochemical, clinical and demographic variables with the plasma concentrations of galectin-3 is shown in Supplementary Table 1 and the variables that showed statistically significant associations are shown in Figs. 2 and 3. A surprising finding of the present study was a negative correlation between galectin-3 concentrations and the number of days that the catheter was indwelling in CVC patients. To understand this association, we verified that there were also negative correlations between the number of days and the circulating concentrations of CRP and procalcitonin (Supplementary Fig. 1), which indicates a decrease in the degree of inflammation, perhaps due to the medication that these patients received.

**Fig 1.**
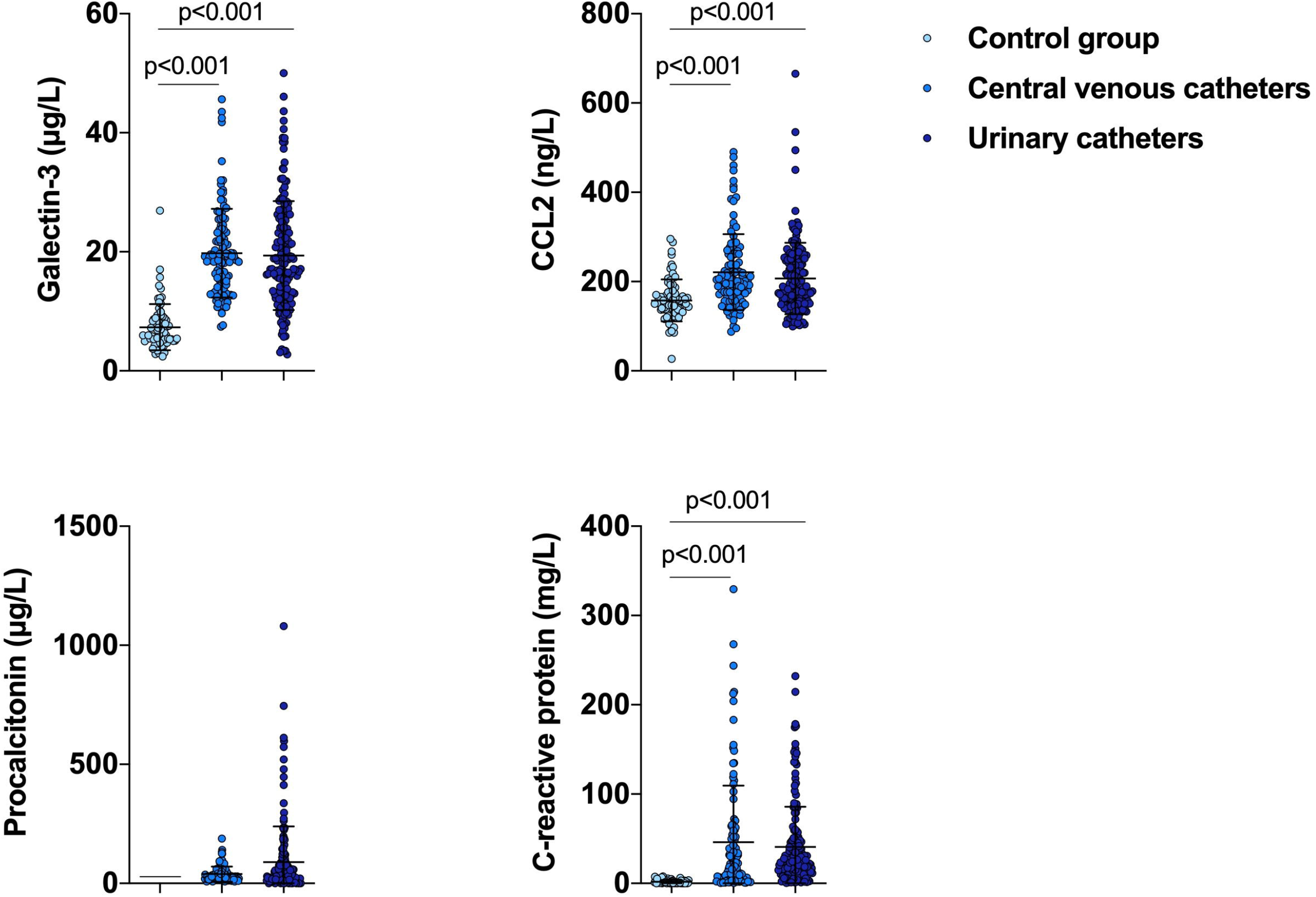
Selected biochemical variables in catheter-bearing patients and the control group. Statistical analyses used the Mann Whitney U test.

**Fig 2.**
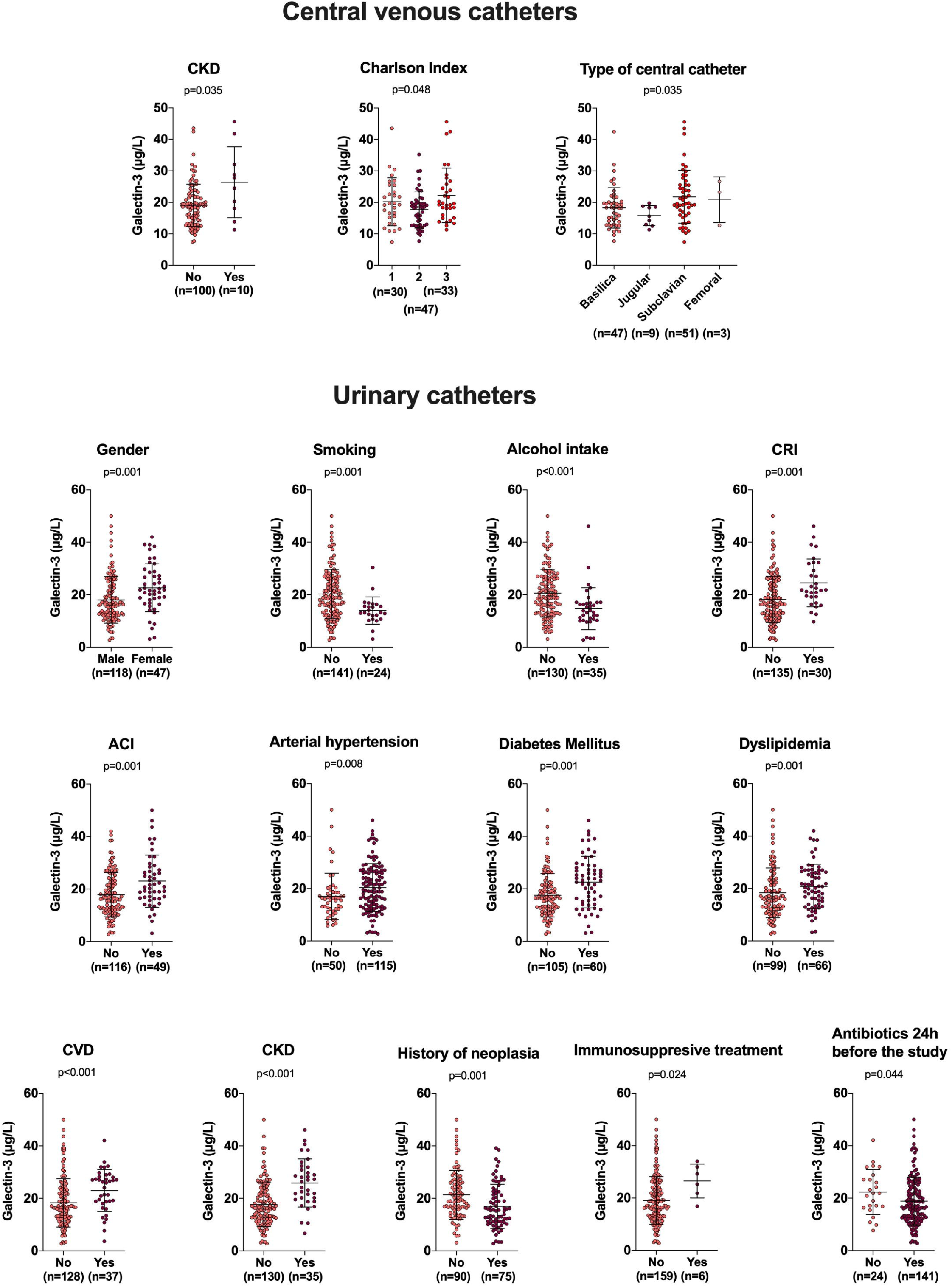
Influence of demographic and clinical variables on plasma galectin-3 concentrations in catheter-bearing patients. Statistical analyses used the χ2 test or the Kruskal-Wallis test.

**Fig 3.**
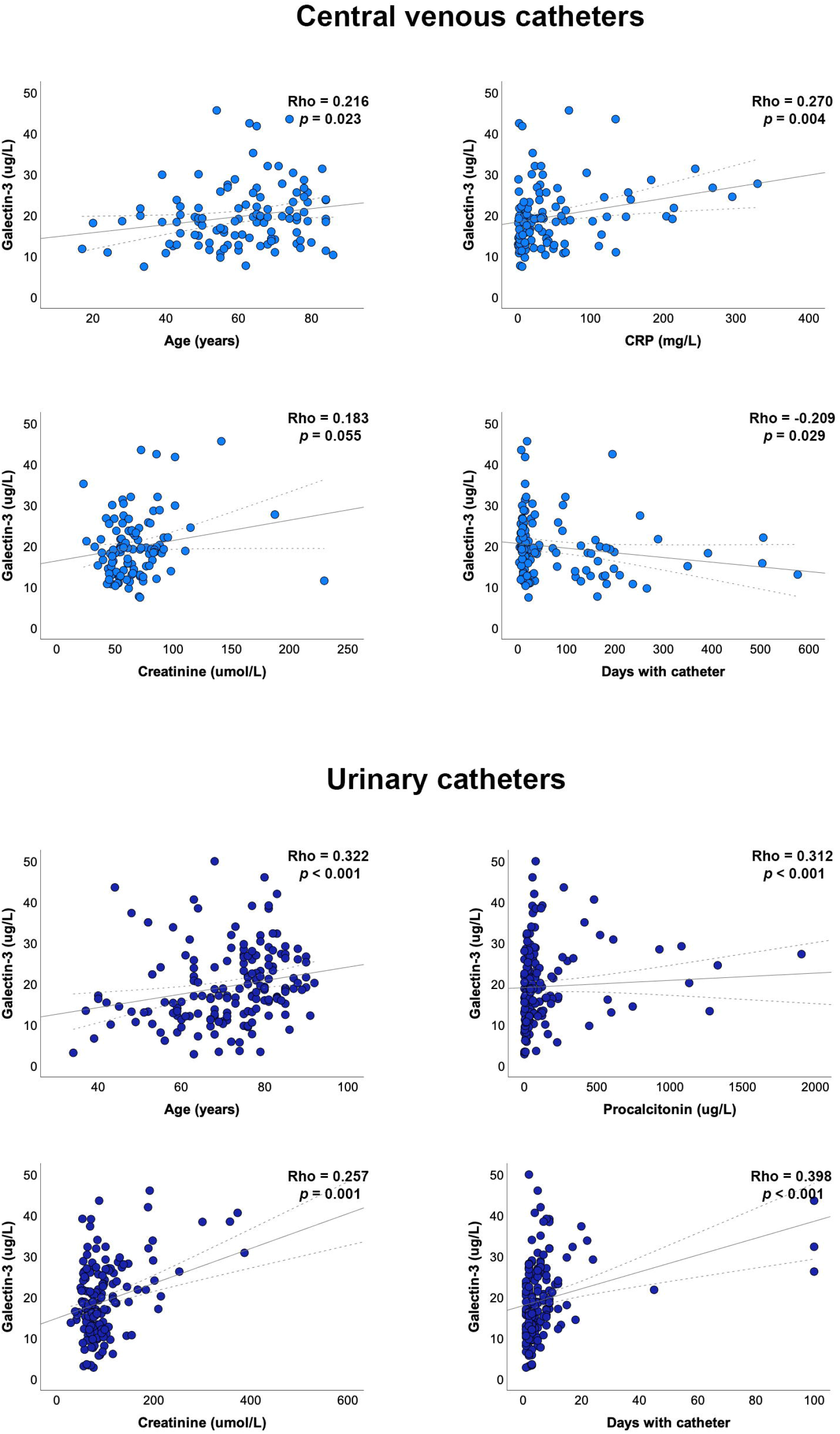
Relationships between selected clinical and biochemical variables and plasma galectin-3 concentrations in catheter-bearing patients. Statistical analyses used Spearman’s Rho test.

Multiple regression analysis showed that the presence of chronic kidney disease was an independent determinant of plasma galectin-3 concentrations in CVC patients (Table 2) and that serum creatinine concentration, cardiovascular disease and the number of days the catheter was indwelling were independent determinants of plasma galectin-3 concentrations in UC patients (Table 3).

**Table 2.**
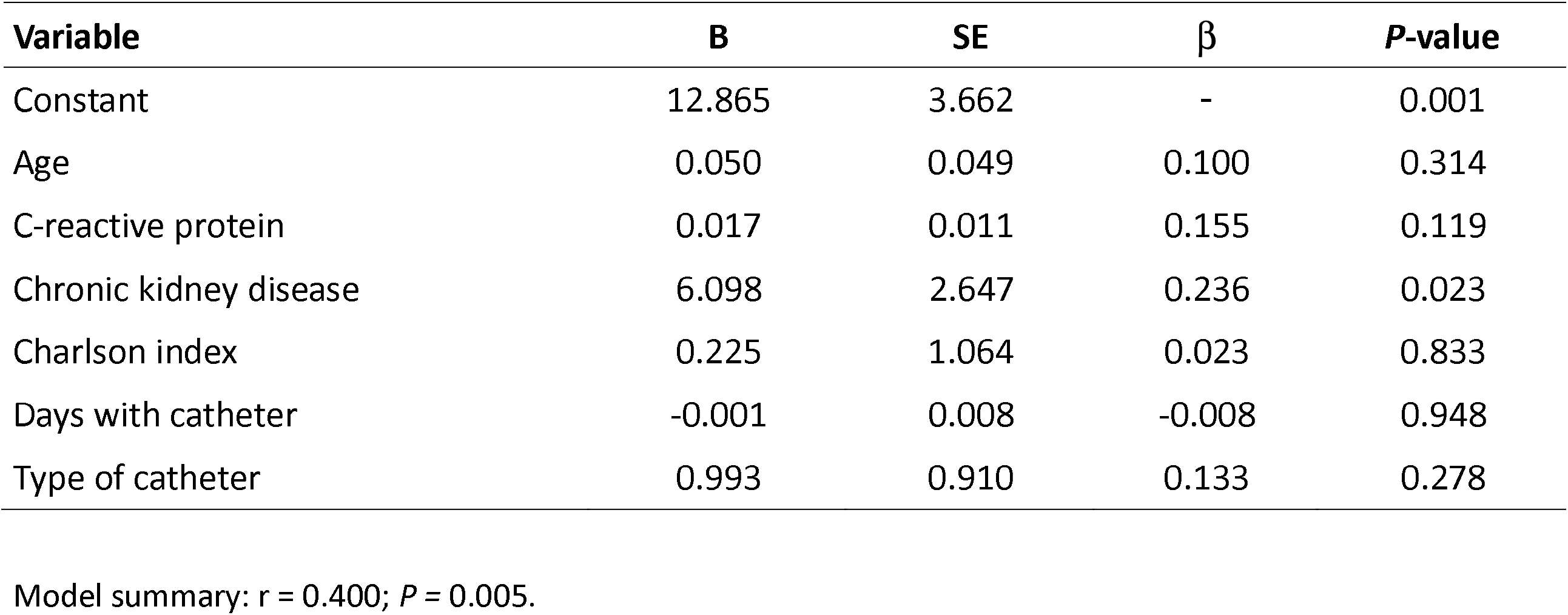
Multiple regression analysis of the combined influence of clinical characteristics and biochemical variables on plasma galectin-3 concentrations in patients with a central venous catheter.

Finally, we wanted to explore the possibility that galectin-3 determination was a reliable index for the diagnosis of CRI and to compare it with the effectiveness of CRP, prolactin and CCL2 determinations. When comparing the diagnostic accuracy of the ROC curves, we found that the order of effectiveness was CRP > CCL2 > procalcitonin > galectin-3 in patients with a CVC, and galectin-3 > CRP ≈ procalcitonin ≈ CCL2 in patients with UC, the area under the curve of galectin-3 being far higher than that of the other parameters in these patients (Fig. 4). At galectin-3 = 15 ng/mL, sensitivity was 90% and specificity was 41%. Values below 15 ng/mL were very unlikely to indicate CRI.

**Fig 4.**
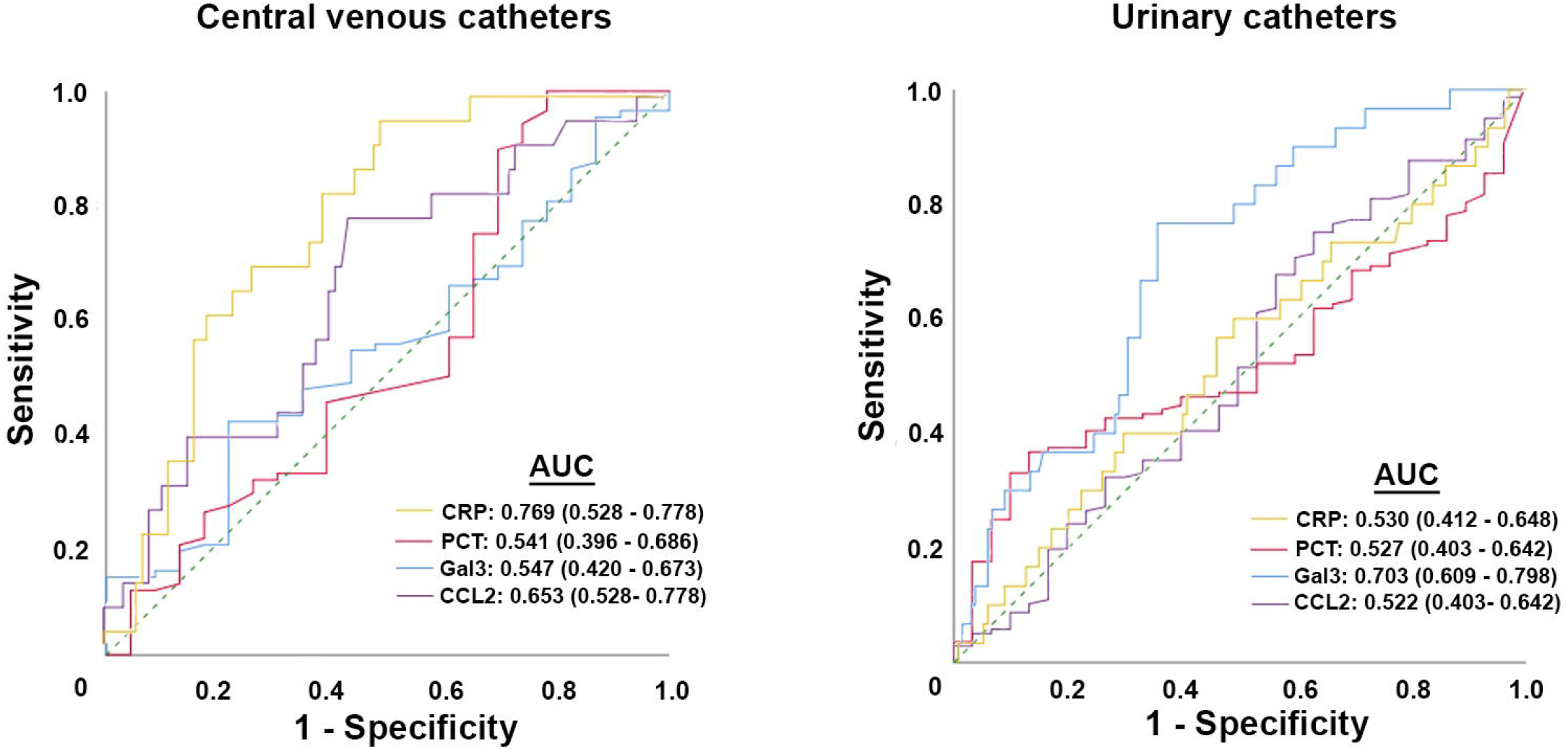
Receiver operating characteristics (ROC) plot of selected biochemical variables in catheter-bearing patients with respect to presentation of catheter-related infection. AUC: area under the curve; CRP: C-reactive protein; Gal3: Galectin-3; PCT: Procalcitonin.

## 4. Discussion

Our results show that catheter-bearing patients have higher plasma galectin-3 concentrations than those of the healthy population. The increase in the levels of this protein is very similar to that of the concentrations of CRP, procalcitonin and CCL2 and they present significant correlations, which suggests that the increase in the concentrations of galectin-3 is related to the inflammatory processes subsequent to the diseases of these patients. The pathologies underlying the use of catheters are diverse and the factors that can influence the increase in galectin-3 are therefore multiple. Indeed, the mere use of catheters can produce an inflammatory reaction. Catheters are made from a variety of materials combined with different chemicals, and it seems as if these chemical substances can dissolve from the catheter material, causing an inflammatory reaction [25,26]. However, the factors that influence galectin-3 concentrations in our study are different in patients with CVC or UC. In the former, high levels of galectin-3 are associated with the presence of chronic kidney disease, age, and serum CRP concentrations. Patients with higher Charlson index values show slightly higher galectin-3 values, indicating a possible effect of associated comorbidities. Likewise, the levels of this protein are also somewhat higher in patients in whom the catheter was placed in the subclavian vein. On the other hand, in patients with a UC, galectin-3 concentrations were associated with multiple factors, including demographic variables, several associated diseases, treatments, and circulating concentrations of procalcitonin and creatinine. If we look for factors common to all types of catheter, we can define age, the level of inflammation (identified by correlations with CRP or procalcitonin), and renal function (identified by the presence of chronic kidney disease or by serum creatinine levels) as common to both CVC and UC patients. Several studies have shown that the plasma concentration of galectin-3 is strongly dependent on renal function. The levels of this protein correlate with glomerular filtration rate in patients with cirrhosis of the liver [27] or polycystic ovary [28]. High concentrations of galectin-3 in plasma have been associated with a risk of chronic kidney disease, and are reduced by hemodialysis treatment [29-31]. Taken together, our data and those of reports by other authors indicate that renal function is an important determinant of the circulating levels of galectin-3.

We found a significant positive association between galectin-3 concentrations and the number of days the catheter was indwelling in patients with UC. This finding might have considerable clinical relevance since long term localization of catheters *in situ* can result in the formation of bacterial biofilms that become resistant to antibiotic treatments [32]. Catheter insertion usually provokes inflammation, which can, in turn, result in hampered antimicrobial capacity of the host immunity due to the effort of immune cells being directed to degrade the foreign material. Ineffective clearance by immune cells is a perfect opportunity for bacteria to attach and form a biofilm [33]. The more days the catheter is worn, the more opportunities the bacteria have to form a biofilm and increase the inflammatory reaction [34]. Biofilms can be developed as a result of physico-chemical interactions between microorganisms and the surface of the supporting catheter, together with specific greater adhesions found on the cell surface or filamentous structures of the bacterial wall, such as *pili* and *fimbriae*. Once attached, bacteria synthesize an extracellular polymeric matrix that provides a structural support for the colony. The outcome is antimicrobial resistance, either because of the physical barrier to the entry of therapeutic drugs, or the difficulty the antibacterial agents encounter in diffusing into the biofilm [35].

Perhaps one of the most important findings is that the measurement of plasma galectin-3 concentrations can discriminate with a high sensitivity -although with a low specificity-between UC patients who have CRI and those who do not. This finding suggests that the determination of plasma galectin-3 may be an effective addition to the laboratory’s panel of tests for the diagnosis of CRI in patients with UC. That may be important for decision-making regarding the treatment of patients with a urinary tract infection. In patients with a suspected urinary tract infection, blood and urine tests, hemocultures, and radiographs should be routinely carried out and clinicians can then diagnose and treat based on the results. Monitoring the infections of an elderly person or hospitalized patient (intubated, postoperative, sedated, etc.) using an appropriate biomarker will help to make decisions sooner, with the consequent benefit for the patient and the reduced economic expense of complementary examinations. A combined sensitivity of 90% with a specificity of 40% results in a saving of about 1/3 of urine bacterial cultures from catheterized patients, leaving 2/3 for outcome from culturing. Therefore, the usefulness of determining galectin-3 as a marker of CRI is limited, but taking into account the enormous number of patients with suspected urinary tract infection in hospitals and residences for the elderly, its implementation in clinical practice would result in relevant improvements in the treatment of patients and in the reduction of expenses. To-date, urine culturing is the gold standard method, but requires considerable time [36]. Simpler, more rapid, urinary dipstick analysis and microscopic examination of urinary sediment are not adequate for the diagnosis of urinary tract infection because of their lack of sensitivity [37]. Hence, the quest for the identification of appropriate biomarkers for a rapid and correct diagnosis of CRI is of considerable value. Very good results have been reported recently for the Sysmex UF-5000 fluorescence flow cytometer for urianalysis, with a very high sensitivity and specificity for the diagnosis of CRI [38], but we think that galectin-3 determination may be an interesting alternative for those laboratories that do not have this technology. In this regard, galectin-3 measurement can be a highly-sensitive parameter for the identification of patients with CRI and is easy and inexpensive to carry out.

## 5. Limitations of the study

We are aware that our study has several limitations. First, the patient groups are heterogeneous. The medical reasons that determine the catheterization of a patient are diverse and the underlying pathologies are multiple. There are also a variety of microbial agents that can infect catheters. However, this diversity reflects the usual clinical practice faced by the physician and, regardless of the cause, the metabolic reactions produced, oxidative stress and inflammation, are similar. In addition, we have analyzed galectin-3 levels through a manual ELISA, which is not very practical for a clinical laboratory that must deliver results quickly. However, galectin-3 assays have been reported in automated analyzers, such as the VIDAS [39] or the ARCHITECT [40], with good results and excellent correlations (r > 0.90) with the manual ELISA, which is considered the reference method.

## 6. Conclusion

The determination of the plasma concentration of galectin-3 may be a useful parameter for assessing the inflammatory status of catheter-bearing patients and may contribute to the diagnosis of CRI in those with a UC.

## Data Availability

Data are available from the corresponding author upon reasonable request.

## Acknowledgments

Editorial assistance was provided by Phil Hoddy at the Service of Linguistic Resources of the Universitat Rovira i Virgili.

## Appendix A. Supplementary data

Supplementary data to this article can be found online at

## Abbreviations

ACI: acute concomitant infection
CCL2: chemokine (C-C) motif ligand 2
CRI: catheterrelated infection
CRP: C-reactive protein
CVC: central venous catheter
UC: urinary catheter
ROC: receiver operating characteristics.

