## Supplementary materials for "Factors influencing plasma galectin-3 concentrations in catheter-bearing hospitalized patients"

**Supplementary Table 1**

Correlations between plasma galectin-3 concentrations and age, number of days the catheter was indwelling, and selected biochemical variables in all study groups.

|  | **Control group**  **(*n* = 72)** | **Central venous catheter**  **(*n* = 110)** | **Urinary catheter**  **(**n **= 165)** |
| --- | --- | --- | --- |
| Age | ρ = 0.203; *P* = 0.087 | ρ = 0.216; *P* = 0.023 | ρ = 0.322; *P* < 0.001 |
| Procalcitonin | <DL | ρ = 0.133; *P* = 0.165 | ρ = 0.312; *P* < 0.001 |
| C-reactive protein | ρ = 0.134; *P* = 0.263 | ρ = 0.270; *P* = 0.004 | ρ = 0.053; *P* = 0.497 |
| Chemokine (C-C motif) ligand 2 | ρ = 0.091; *P* = 0.445 | ρ = 0.111; *P* = 0.248 | ρ = 0.027; *P* = 0.734 |
| Creatinine | ρ = -0.251; *P* = 0.330 | ρ = 0.183; *P* = 0.055 | ρ = 0.257; *P* = 0.001 |
| Days with catheter | NA | ρ = -0.209; *P* = 0.029 | ρ = 0.398; *P* < 0.001 |

CVC: Central venous catheter; <DL: Below the detection limit of the assay; UC: Urinary catheter; NA: Not applicable. Correlations were calculated with the Spearman’s ρ test.

**Supplementary Table 2**

Relationships between plasma galectin-3 concentrations and clinical and demographic characteristics in all study groups^a^.

|  | **Control group**  **(*n* = 72)** | **Central venous catheter**  **(*n* = 110)** | **Urinary catheter**  **(*n* = 165)** |
| --- | --- | --- | --- |
| Gender, male | 0.206 | 0.468 | 0.001 |
| Smoking | 0.318 | 0.568 | 0.001 |
| Alcohol habit | 0.442 | 0.586 | < 0.001 |
| Acute concomitant infection | NA | 0.179 | 0.001 |
| Type of microorganism^b^ | NA | 0.565 | 0.150 |
| Type of central catheter^c^ | NA | 0.035 | NA |
| Catheter-related infection | NA | 0.492 | 0.001 |
| Antibiotics treatment | NA | 0.986 | 0.044 |
| Charlson index | NA | 0.048 | 0.190 |
| Mc Cabe index | NA | 0.320 | 0.443 |
| Arterial hypertension | NA | 0.210 | 0.008 |
| Diabetes mellitus | NA | 0.057 | 0.001 |
| Dyslipidemia | NA | 0.679 | 0.025 |
| Cardiovascular disease | NA | 0.195 | < 0.001 |
| Chronic kidney disease | NA | 0.035 | < 0.001 |
| Chronic neurovascular disease | NA | 0.236 | 0.325 |
| Chronic obstructive pulmonary disease | NA | 0.608 | 0.840 |
| History of cancer | NA | 0.135 | 0.001 |
| Immunosuppressive treatment | NA | 0.142 | 0.024 |

^a^ This table represents the *p* values comparing galectin-3 concentrations in subjects with a certain characteristic with those who do not, using the Mann Whitney *U* test (two independent samples) or the Kruskal-Wallis test (more than two independent samples). ^b^ Gram-positive, Gram-negative or funghi. ^c^ Place of central catheter insertion: Basilica, jugular, subclavian or femoral. NA: Not applicable.

**Supplementary Fig. 1**

Relationships between the number of days the catheter was indwelling and the circulating levels of procalcitonin, C-reactive protein (CRP) and the chemokine (C-C) motif ligand 2 (CCL2) in patients with a central venous catheter.

00

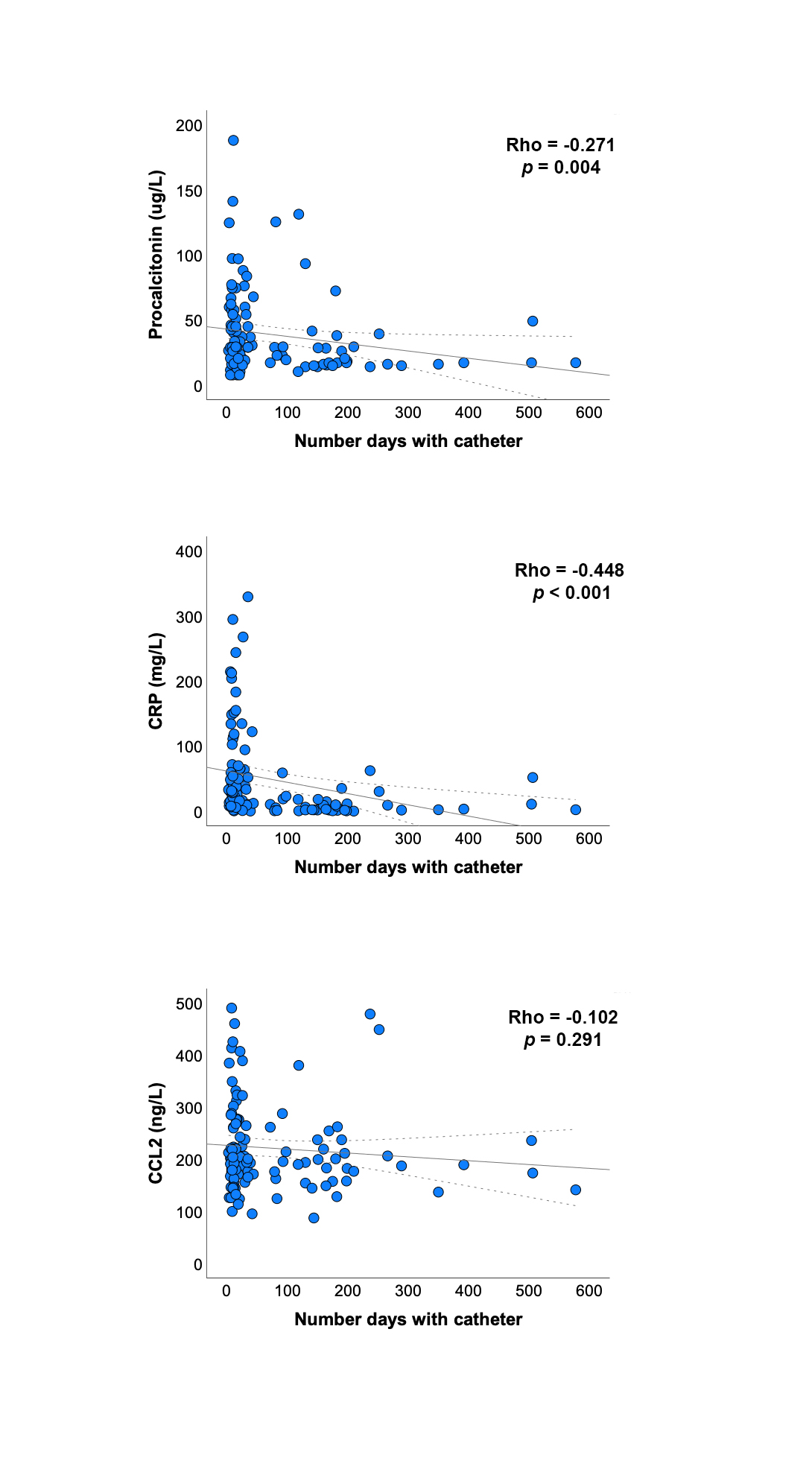
